# Spatiotemporal Analysis of COVID-19 Risk in Malawi: The Impact of Age, Poverty, Population Density, and Environmental Factors

**DOI:** 10.1101/2025.02.16.25322329

**Authors:** Mwandida Kamba Afuleni, Ian Hall, Thomas House, Olatunji Johnson

## Abstract

**Background:** Understanding the spatiotemporal variation of COVID-19 transmission and its determinants is crucial for gaining deeper insights into the dynamics of disease spread. Regional and temporal differences in demographics, socioeconomic conditions, and environmental factors shaped the trajectory of the COVID-19 pandemic, underscoring the importance of advanced spatio-temporal modelling. This research aims to construct a spatiotemporal model to examine the relationship between age groups, poverty, population density, precipitation, and COVID-19 risk, as well as to pinpoint high-risk areas.

**Methods:** Here we present a spatiotemporal statistical analysis using COVID-19 case data from Malawi recorded from 2 April 2020 to 27 March 2022. Bayesian spatiotemporal models were fitted, with weekly confirmed cases as the response variable and demographic, socioeconomic, and environmental factors as predictors.

**Results:** The findings reveal that spatial and temporal factors, along with age, population density, and poverty, significantly affect observed COVID-19 incidence in Malawi, whereas precipitation does not. The greatest risk was observed during colder months (June–July), December’s festive season, and January. Urban centres and lake-shore districts were disproportionately impacted, with individuals aged 40 − 49 at particularly high risk.

**Conclusions:** These results emphasise the need to prioritise vaccinations for working-age populations in urban and tourist areas during high-risk periods. Moreover, ensuring adherence to public health guidelines and enhancing healthcare services in these districts is critical.

**Key Messages:**

- The age group 40–49 faced a significantly higher risk of COVID-19 compared to all other age groups.
- COVID-19 risk is generally low across Malawi during the study period, with Blantyre being a notable exception.
- Positive associations were identified between COVID-19 risk and factors such as age, poverty levels, and population density.
- Both spatial and temporal dynamics were found to have substantial impacts on COVID-19 transmission risk.

## 1. Introduction

Infectious diseases remain a significant global concern and leading cause of death [1]. Over the past two decades, respiratory infectious diseases caused by coronaviruses like the Severe Acute Respiratory Syndrome Coronavirus (SARS-CoV), the Middle East Respiratory Syndrome Coronavirus (MERS-CoV), and Severe Acute Respiratory Syndrome Coronavirus-2 (SARS-CoV-2) have posed major threats to global health [2, 3]. SARS-CoV-2, responsible for COVID-19, emerged in Wuhan, China, in December 2019 and spread worldwide within three months [4, 5]. Declared a Public Health Emergency of International Concern (PHEIC) by the WHO on January 30, 2020 [6], it had infected over 100 million people and caused two million deaths within a year [7]. Africa reported its first case on February 14, 2020, with the virus spreading continent-wide in three months [7]. In Malawi, the first confirmed COVID-19 case was reported in Lilongwe on April 2, 2020, two weeks after the president pre-emptively declared a state of national disaster on March 20 [8, 9, 10].

A range of demographic, socioeconomic, and environmental factors influence SARS-CoV-2 dynamics [11, 12, 13]. Key drivers include age [14], with older individuals (particularly those aged 60 and over) at higher risk [15], and poverty, which is linked to increased risk due to reduced adherence to interventions by low-income populations prioritising daily survival [16, 17]. In the USA, poverty was positively associated with infection risk early in the pandemic however, the association later changed to negative assumably due to case under-ascertainment among the less privileged individuals, when the testing resources became scarce [18]. Population density has also been claimed to impact SARS-CoV-2 risk [13, 15, 16], though some studies found it not significant [19]. Tourism correlates with case surges, as imported cases often contribute to the spread [13]. COVID-19 transmission occurs through respiratory fluids [4, 5, 20], with low temperatures, precipitation and humidity enhancing spread [11, 21, 22, 23]. Air pollution has been linked to higher mortality, with studies reporting increased COVID-19 deaths in heavily polluted areas [12].

Studies on spatial and spatiotemporal modelling of COVID-19 have been conducted in many parts of the world, including Africa. Researchers use geographical information systems (GIS) and / or Bayesian statistical models for analysis [24]. Some studies in Africa include Gayawan *et al.* (2020) [25], who used a two-parameter hurdle Poisson model; Tong *et al.* (2022) [26] who employed GIS; Adekunle *et al.* (2020) [27], who applied generalised additive models; and Abdul (2020) [28], who utilised an endemic-epidemic multivariate time-series model.

At the time of this study, two spatiotemporal modelling studies on SARS-CoV-2 in Malawi had been conducted. One, by Chinkaka *et al.* (2023)[15], covered the period from April 2020 to May 2021 using GIS and found significant effects of age (particularly being 60+ years old) and population density on disease risk. The other, by Ngwira *et al.* (2021)[16], focused on the early phase of the pandemic, when there was only one wave. Conducted from April to October 2020, it used semiparametric spatiotemporal models and found a higher risk of COVID-19 in major cities compared to rural areas, attributing this to higher population density in urban settings. The study also identified significant effects of location, time, and space-time interaction on COVID-19. During this phase, the pandemic peaked in August 2020, with a positive and significant correlation between risk and age (here being 65+ years old).

This study aims to develop a multi-variable spatiotemporal model for SARS-CoV-2 cases in Malawi, incorporating spatial, temporal, age and their interactions. Using data collected from April 2, 2020, to March 27, 2022 – covering four major waves – the study has three primary objectives: (1) to identify the combination of demographic, socioeconomic, and environmental factors driving the spatial variation in SARS-CoV-2 cases across Malawian districts; (2) to map the spatial heterogeneity in the relative risk of SARS-CoV-2; and (3) to identify hotspot districts over time across different age groups.

## 2. Methods

### 2.1. Study area

Malawi is a landlocked country bordered by Mozambique to the east, south, and southwest; Tanzania to the north and northeast; and Zambia to the west and northwest. Geographically, it lies between latitudes 9*^◦^* 22’ S and 17*^◦^* 03’ S, and longitudes 33*^◦^* 40’ E and 35*^◦^* 55’ E. With a total surface area of around 120,000 km^2^ [29], Malawi has an estimated population of just over 18 million [30]. The country is divided into three regions—Northern, Central, and Southern—which are further subdivided into 28 districts. According to the 2018 Population and Housing Census, the capital city, Lilongwe, located in the Central Region, has the highest population proportion at 9.3%. Over 80% of the population resides in rural areas, with 16% in urban centres. Notably, half of Malawi’s population is aged 17 years or younger [30]. Fig.1A illustrates a map of Malawi, highlighting its 27 districts on the mainland.

**Figure 1:**
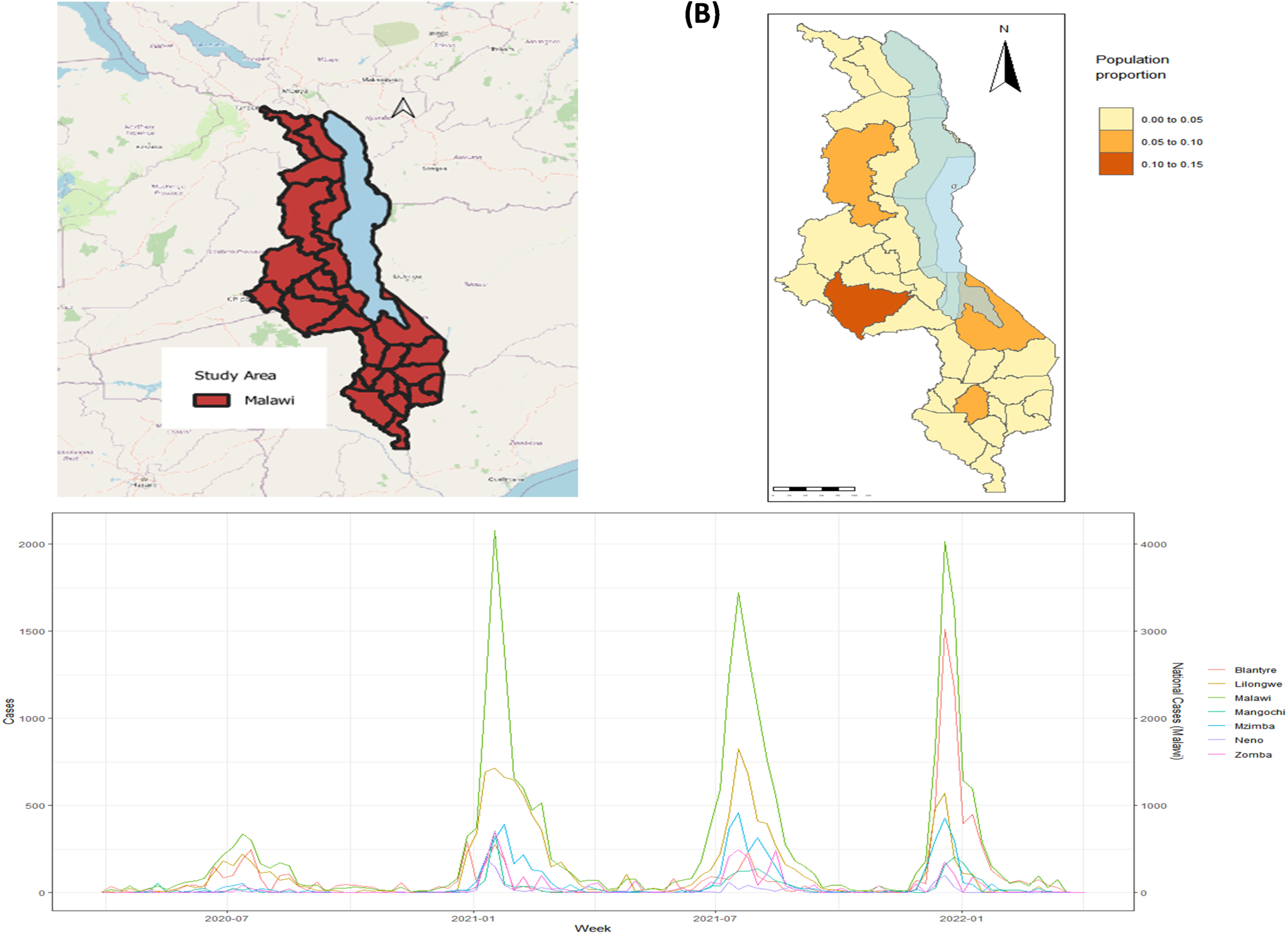
(A) The study area (Malawi). Map was created using QGIS version 3.40.0 − 1 available at https://qgis.org/. (B) Population proportion map. The blue-shaded area is Lake Malawi. The shapefiles for the districts were obtained from https://gadm.org/data.html while the Lake Malawi shapefile was obtained from the Natural Earth website, https://www.naturalearthdata.com/. (C) National and district curves for raw data.

### 2.2. Data and data sources

Line list data on confirmed COVID-19 cases collected from 2 April 2020, to 27 March 2022 was obtained from the Ministry of Health (MoH) through the Public Health Institute of Malawi (PHIM) [31]. This dataset includes information on all individuals who tested positive for COVID-19, detailing variables such as case ID, report date, date of birth, age, gender, district, and other relevant attributes. Population totals and population density for the districts were sourced from reports by the National Statistical Office of Malawi (NSO) available at https://malawi.unfpa.org/en/publications/malawi-2018-population-and-housing-census-main-report [30]. Data on the proportion of the population living on $1.25 or less per day, measured per grid square (1 km by 1 km at the equator), was obtained from the WorldPop website https://hub.worldpop.org/doi/10.5258/SOTON/WP00290 [32]. The poverty proportion for each district was subsequently calculated using a population-weighted average across the district. Environmental data was also considered in this study. In particular, precipitation and humidity data were obtained from NASA POWER Project Team (2024) [33] and temperature data was obtained from WorldClim Team (2024) [34].

### 2.3. Outcome variable

The outcome variable is the weekly number of confirmed COVID-19 cases, starting from the date of the first recorded case on April 2, 2020, to March 27, 2022, encompassing the first four waves over 105 epidemiological weeks. Data from April 2022 onward was excluded from the analysis, as almost all districts reported zero cases, rendering the data unsuitable for modelling. Case data was collected by the Ministry of Health through polymerase chain reaction (PCR) tests conducted on nasal samples from individuals presenting symptoms of the infection and those identified as close contacts of confirmed cases.

### 2.4. Covariates

The variables considered in the model are age, population density, poverty, precipitation, humidity and temperature, as these factors have been shown to influence the risk of COVID-19. Population density is included as a covariate because it has been identified as a factor that might impact on the spread of SARS-CoV-2, given the role of social mixing in transmission [13, 35]. Similarly, poverty is considered due to its reported impact on SARS-CoV-2 transmission [16, 18], though studies differ on the nature of this association. This study seeks to determine whether poverty has a significant effect on COVID-19 risk and to explore whether the correlation is positive or negative. Fig.1B illustrates the total population across districts, while Fig.2A and Fig.2B depict the spatial distribution of key geospatial covariates.

**Figure 2:**
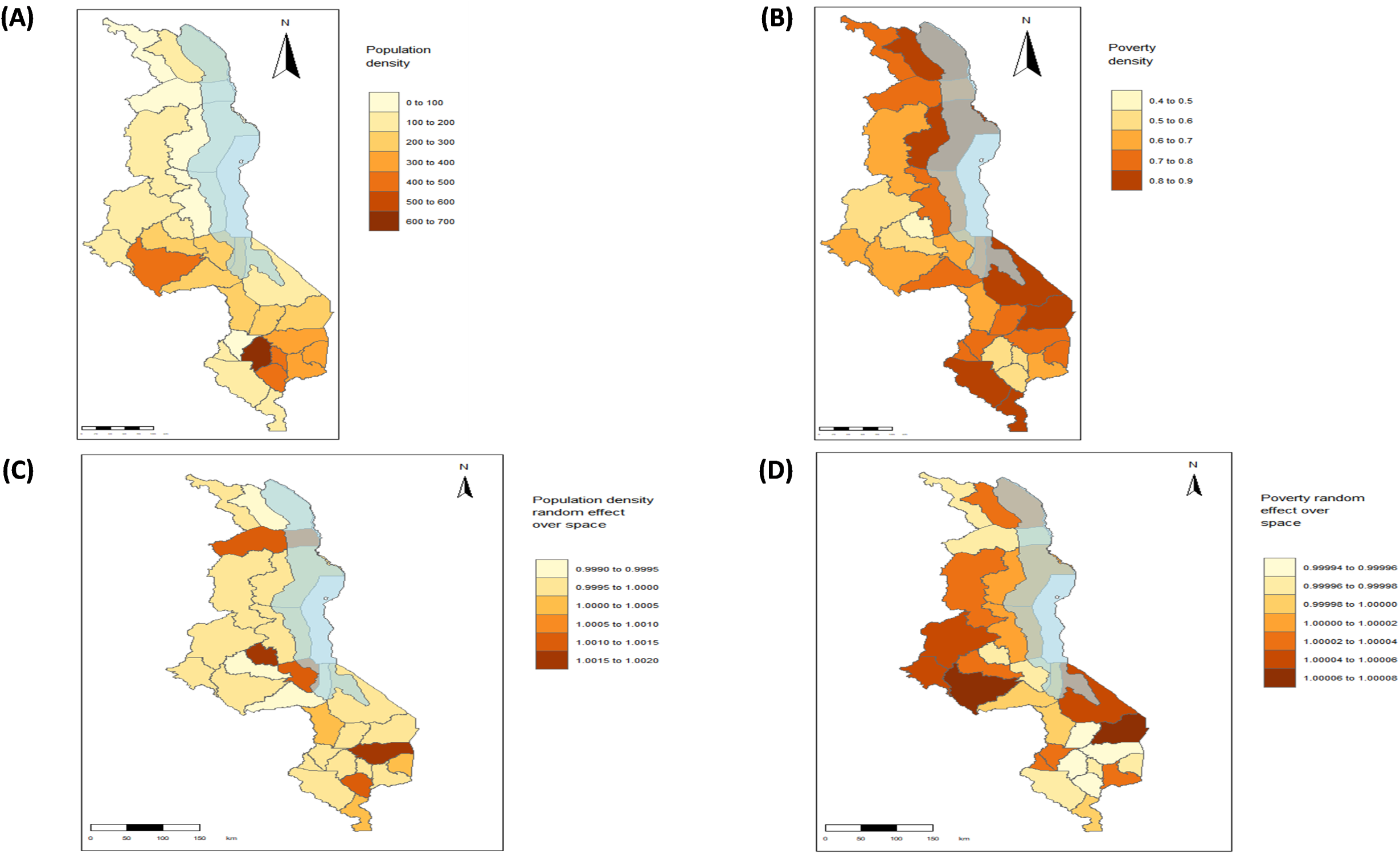
(A) Population density. (B) Poverty density. (C) Effect of population density over space. (D) Effect of poverty over space. The blue-shaded area in the maps is Lake Malawi. The shapefiles for the districts were obtained from https://gadm.org/data.html while the Lake Malawi shapefile was obtained from the Natural Earth website, https://www.naturalearthdata.com/

It is widely recognized that there is a direct association between COVID-19 incidence and old age. While the older age group was included as a predictor in this study, the research was extended also to examine the young and middle-aged groups. We used quantile-based grouping to create five age groups of roughly equal number of cases and this was done for the following reasons: balanced representation, model stability, help reduce noise and facilitate Comparability among groups leading to more interpretable results. The age groups include: 0 − 1, 20 − 29, 30 − 39, 40 − 49 and 50+. Fig. 1 in the Supplementary Material 3.2 shows an age histogram. The mean and median ages are 36.24 and 34, respectively. Additionally, 25% of the study population is 46 years old or above.

Table 1 presents the description and spatial and temporal resolution of the variables used in this study.

**Table 1:** Data sources, description and properties of the variables

| Category | Variable | Variable description and pre-processing | Temporal resolution (weeks) | Temporal coverage | Spatial resolution |
| --- | --- | --- | --- | --- | --- |
| *Outcome* | COVID-19 cases | Weekly number of confirmed COVID-19 cases for the first four waves over 105 epidemiological weeks for each district. | 1 | 2 April 2020 to 27 March 2022. | District level |
| *Demographic and socioeconomic covariates* | Age | Age group of the confirmed cases; 0-19, 20-29, 30-39, 40-49 and 50+. The age brackets were determined using quantiles and demographic data on the age distribution of the total population. | None | 2018 | District level |
|  | Population density | Number of people per km<sup>2</sup>. | None | 2018 | District level |
| | Poverty | The proportion of the population living in poverty, defined as living on under \$1.25 per day, was extracted from a tagged image file (TIF). This data was read into R as a **stars** object, and then aggregated to calculate the poverty proportion for each district. | None | 2010 | 1km by 1km at the equator |
| *Environmental covariates* | Temperature | The quantitative measure of heat present in an environment, influencing physical and biological processes population-weighted averaged over the districts and measured in degrees celsius. | 1 | April 2020 to March 2022 | 1km by 1km |
|  | Relative Humidity | A percentage of the maximum amount of moisture the air can hold at a given temperature, population-weighted averaged over the districts. | 1 | April 2020 to March 2022 | 55.6 km by 55.6 km. |
|  | Precipitation | Any type of water that forms in the Earth's atmosphere and then drops onto the surface of the Earth population-weighted averaged over the districts and measured in millimeters. | 1 | April 2020 to March 2022 | 55.6 km by 55.6 km. |

*2.5. Model formulation*

### 2.5. Model formulation

We used Bayesian hierarchical spatio-temporal framework to model the weekly observed counts of COVID-19 cases. Let *i* = 1, 2*, … ,* 28 denote the index for the spatial areas (districts), *j* = 1, 2*, … ,* 105 denotes the time points in weeks from 2 April 2020 to 31 December 2022 and *k* = 1, 2*, …,* 5 denotes the index for age groups 0-19, 20-29, 30-39, 40-49, and 50+. Let *Y_ijk_* denote the random variable of the number of SARS-CoV-2 confirmed cases in district *i*, week *j* and age category *k*. Then we model *Y_ijk_* as follows:

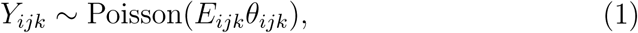

where *E_ijk_* is the expected number of cases in the absence of any heterogeneity in individual risk and *θ_ijk_* is the relative risk (RR) in district *i*, week *j* and age group *k*. *E_ijk_* is therefore calculated as,

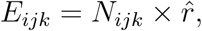

where *N_ijk_*is the population in district *i*, week *j* and age group *k* and

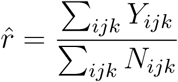

is the global observed disease rate. The log-relative risk is modelled as

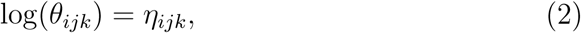

where *η_ijk_* is a linear predictor. This linear predictor is modelled as a linear combination of fixed and random effects and ten candidate models were examined as shown in Table 2, with prior specification in the Table caption.

**Table 2:** Random effects models for district *i*, time *j* and age *k*. 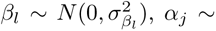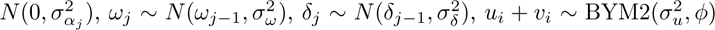, three structures for linear time trend 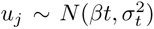, Random walk 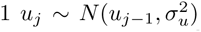, Random walk 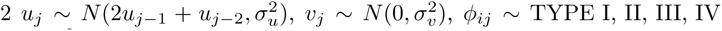 [36] and 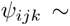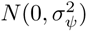

| Model | Linear predictor $\eta_{ijk}$ |
| --- | --- |
| I | $\beta_0 + \alpha_k + \beta_1 \text{PRE}_{ijk} + \omega_j + \delta_j$ |
| II | $\beta_0 + \alpha_k + \beta_1 \text{PRE}_{ijk} + \omega_j + \delta_j + u_i + v_i$ |
| III | $\beta_0 + \alpha_k + \beta_1 \text{PRE}_{ijk} + \omega_j + \delta_j + u_i + v_i + u_j + v_j$ |
| IV | $\beta_0 + \alpha_k + \beta_1 \text{PRE}_{ijk} + \omega_j + \delta_j + u_i + v_i + u_j + v_j$ |
| V | $\beta_0 + \alpha_k + \beta_1 \text{PRE}_{ijk} + \omega_j + \delta_j + u_i + v_i + u_j + v_j$ |
| VI | $\beta_0 + \alpha_k + \beta_1 \text{PRE}_{ijk} + \omega_j + \delta_j + u_i + v_i + u_j + v_j + \phi_{ij}$ |
| VII | $\beta_0 + \alpha_k + \beta_1 \text{PRE}_{ijk} + \omega_j + \delta_j + u_i + v_i + u_j + v_j + \phi_{ij}$ |
| VIII | $\beta_0 + \alpha_k + \beta_1 \text{PRE}_{ijk} + \omega_j + \delta_j + u_i + v_i + u_j + v_j + \phi_{ij}$ |
| IX | $\beta_0 + \alpha_k + \beta_1 \text{PRE}_{ijk} + \omega_j + \delta_j + u_i + v_i + u_j + v_j + \phi_{ij}$ |
| X | $\beta_0 + \alpha_k + \beta_1 \text{PRE}_{ijk} + \omega_j + \delta_j + u_i + v_i + u_j + v_j + \phi_{ij} + \psi_{ijk}$ |

The model specification for the fixed effects is as follows: *β*_0_ is the global intercept; *α_k_* is the age categories specific intercept for category *k*; *β*_1_ is the effect of precipitation (PRE); *ω_j_* is the time-varying effect of poverty; *δ_j_* is the time-varying effect of population density. The spatial random effects, represented as *u_i_* + *v_i_*, are modelled using the BYM2 framework [37], an alternative parameterization of the BYM (Besag, York and Mollie) model [38]. This formulation introduces a mixing parameter, *ϕ*, to effectively balance the contributions of *u_i_* (structured spatial effects) and *v_i_* (unstructured spatial effects). *u_i_* is modelled using a conditional autoregressive (CAR) structure [39] and accounts for the spatial dependence between relative risks. This structure assumes that neighbouring areas are more likely to show similar risks than areas that are far apart. On the other hand, *v_i_* models unstructured spatial effects accounting for independent noise, as neighbouring areas can be independent. To capture temporal random effects, the term *u_j_* + *v_j_* is added, where *u_j_* accounts for the correlated temporal random effects, as-suming similar risks at close time points, and *v_j_* models temporal effects that are independent. In the model, *ϕ_ij_* is the space-time interaction term to account for variation that can not be explained by space and time [40]. As shown in Table 2, four types of interactions, resulting from all possible combinations of structured and unstructured spatial and temporal effects, are considered [36], including: Type I interaction, which has an identically independent distribution and represents the interaction of unstructured spatial and unstructured temporal random effects; Type II interaction, which assumes structured temporal effects for each area, independent of all other areas; Type III interaction, which accounts for structured spatial effects for each time unit, independent of all other time points; and Type IV interaction, which captures correlated spatial and temporal effects. Finally, *ψ_ijk_*, representing a space-time-age interaction, is added to extend the four types of space-time interaction. The full priors specification for the models are presented in the Supplementary Material.

### 2.6. Inference and model selection

Analysis was done in R using the INLA package [41]. INLA provides an approximate Bayesian inference framework for latent Gaussian models. It was chosen over Markov Chain Monte Carlo (MCMC) due to its faster computation and ability to efficiently handle large datasets. This approach uses analytical approximations and numerical integration to estimate posterior distributions for model parameters. Model selection was based on the scores of Deviance Information Criterion (DIC), Watanabe-Akaike Information Criterion (WAIC) and the effective number of parameters.

## 3. Results

### 3.1. Understanding case trends in the districts

Nationwide and district-level trends for confirmed cases over the study period are depicted in Fig. 1C. Higher case counts were observed in major urban centres, including Blantyre, Lilongwe, Mzimba/Mzuzu, and Zomba, as well as in Neno and Mangochi. A sharp rise in cases occurred in Blantyre during the fourth wave, likely linked to the characteristics of the Omicron variant, which dominated this wave. Previous studies on the epidemiological and phylogenetic analyses of SARS-CoV-2 indicated that Omicron prevalence was notably higher in southern Malawi, where Blantyre is located. For improved comparison across districts, the data were standardized, with the results presented in Fig. S2 of the Supplementary Material 3.2. Standardised case numbers were exceptionally high in Neno during the second wave and in Blantyre during the fourth wave. Additionally, Zomba, Lilongwe, Mzuzu, and Mangochi experienced elevated risks at various points compared to other districts not explicitly highlighted.

### 3.2. Model fitting

Before fitting spatiotemporal models, a Poisson generalised linear model (GLM) with explanatory variables age, poverty, population density, precipitation, humidity, and temperature, was fitted to the data. Interestingly, the summary for the GLM showed that risk was high in the age group 40-49, followed by the age group 50+. The minimal risk was observed in age group 0-19. Additionally, we found population density (*δ* = 0.0019, 95% CRI (0.0018, 0.0019)) and poverty (*ω* = 0.0325, 95% CRI (0.0209, 0.0440)) to be significant. Furthermore, we found that out of the environmental variables, precipitation was significant, therefore in subsequent models, we only use precipitation in our model.

Then we fitted models I to X as summarized in Table 2 and the model with the least DIC and WAIC scores as well as the moderate effective number of parameters was selected as the best fit (see Table 3).

**Table 3:**
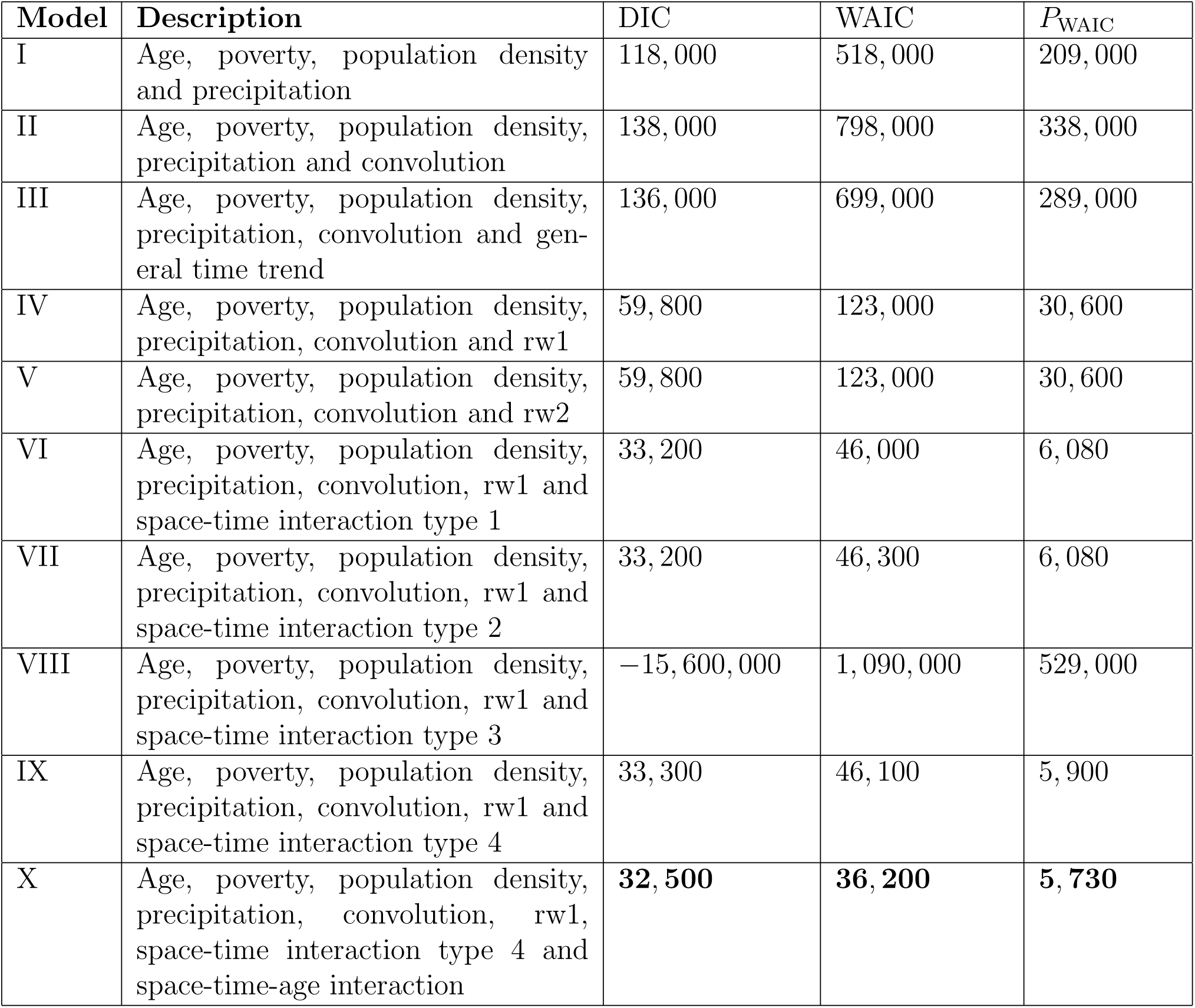
Table showing the result of the model selection criteria (DIC, WAIC and effective number of parameters for WAIC) for the ten models considered.

| Model | Description | DIC | WAIC | $P_{\text{WAIC}}$ |
| --- | --- | --- | --- | --- |
| I | Age, poverty, population density and precipitation | 118,000 | 518,000 | 209,000 |
| II | Age, poverty, population density, precipitation and convolution | 138,000 | 798,000 | 338,000 |
| III | Age, poverty, population density, precipitation, convolution and general time trend | 136,000 | 699,000 | 289,000 |
| IV | Age, poverty, population density, precipitation, convolution and rw1 | 59,800 | 123,000 | 30,600 |
| V | Age, poverty, population density, precipitation, convolution and rw2 | 59,800 | 123,000 | 30,600 |
| VI | Age, poverty, population density, precipitation, convolution, rw1 and space-time interaction type 1 | 33,200 | 46,000 | 6,080 |
| VII | Age, poverty, population density, precipitation, convolution, rw1 and space-time interaction type 2 | 33,200 | 46,300 | 6,080 |
| VIII | Age, poverty, population density, precipitation, convolution, rw1 and space-time interaction type 3 | -15,600,000 | 1,090,000 | 529,000 |
| IX | Age, poverty, population density, precipitation, convolution, rw1 and space-time interaction type 4 | 33,300 | 46,100 | 5,900 |
| X | Age, poverty, population density, precipitation, convolution, rw1, space-time interaction type 4 and space-time-age interaction | **32,500** | **36,200** | **5,730** |

Model VIII has an extremely large negative DIC and a high WAIC, indicating poor data fit or excessive model complexity, which renders its estimates unreliable. The analysis proceeded with the best-fitting model, Model X, which includes age and precipitation as fixed effects, along with poverty and population density as random effects. The model also incorporates spatial, temporal, and spatiotemporal interaction of type 4, as well as an independent space-time-age interaction. As shown in Table 4, for the fixed effects, individuals aged 40-49 are at the highest risk, followed by those in the oldest age group (50+). The elevated risk in the 40-49 age group is likely due to their status as the most productive and active working demographic. A positive correlation is observed between COVID-19 risk and precipitation; however, this relationship is not statistically significant as the confidence interval includes zero. All random effects, including poverty, population density, spatial, temporal, and the interactions of space and time, as well as space, time, and age, show significant effects on COVID-19 risk, as indicated in Table 4.

**Table 4:**
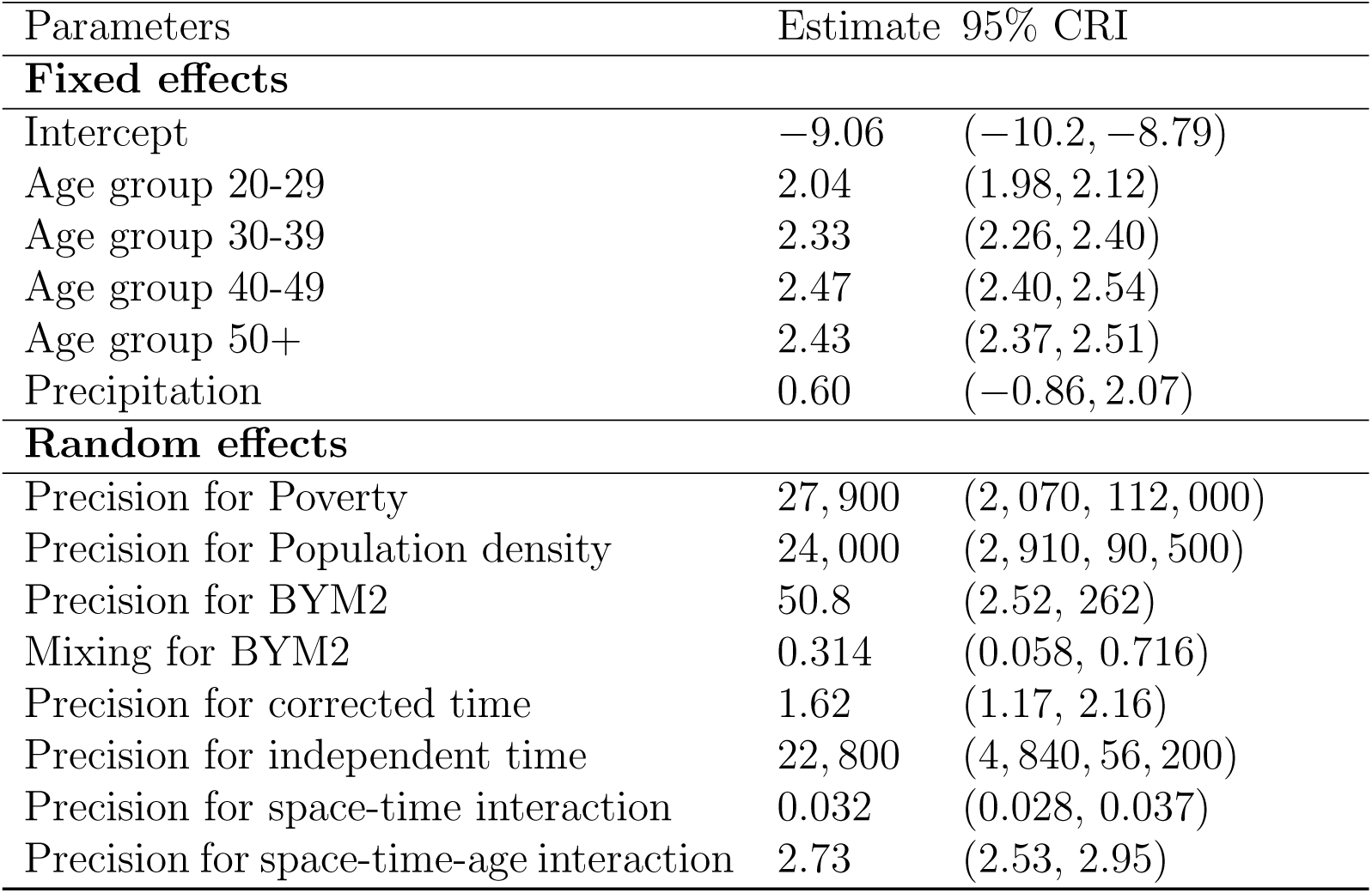
Summary of the parameter estimate (posterior mean) of the fixed and random effects for model X and their corresponding 95% credible interval (CRI)

| Parameters | Estimate | 95% CRI |
| --- | --- | --- |
| **Fixed effects** |  |  |
| Intercept | -9.06 | (-10.2, -8.79) |
| Age group 20-29 | 2.04 | (1.98, 2.12) |
| Age group 30-39 | 2.33 | (2.26, 2.40) |
| Age group 40-49 | 2.47 | (2.40, 2.54) |
| Age group 50+ | 2.43 | (2.37, 2.51) |
| Precipitation | 0.60 | (-0.86, 2.07) |
| **Random effects** |  |  |
| Precision for Poverty | 27,900 | (2,070, 112,000) |
| Precision for Population density | 24,000 | (2,910, 90,500) |
| Precision for BYM2 | 50.8 | (2.52, 262) |
| Mixing for BYM2 | 0.314 | (0.058, 0.716) |
| Precision for corrected time | 1.62 | (1.17, 2.16) |
| Precision for independent time | 22,800 | (4,840, 56,200) |
| Precision for space-time interaction | 0.032 | (0.028, 0.037) |
| Precision for space-time-age interaction | 2.73 | (2.53, 2.95) |

Further analysis of the coefficients (log risks) for the age groups, illustrated in the forest plot in the Supplementary Material, reveals that the risk in all age groups is lower than the baseline assumed by the model, typically 1. Additionally, the credible intervals for all groups are narrow and do not cross the zero line, indicating high confidence in the reduced risk. The coefficient for the 0-19 age group is the furthest to the left, suggesting a significantly lower risk compared to the other groups.

Population density and poverty were modelled as random effects. The results showed district-level variations in how population density influenced COVID-19 risk, attributed to unknown factors (see Fig. 2C). In the Southern region, population density in Blantyre and the areas east of Blantyre excluding Thyolo and Zomba was found to contribute less to risk. Similarly, the high population density in Lilongwe city (central region) also had a minimal contribution to COVID-19 risk. These findings imply that other unmeasured factors may be driving risk patterns in these urban centers. Notably, districts such as Zomba city and Thyolo (South), Salima and Ntchisi (Central), and Rumphi (North) displayed higher-than-expected COVID-19 risk. This suggests that unknown factors associated with population density, such as mobility patterns, densely populated living spaces or compliance with risk prevention measures, could be influencing these variations. The analysis also revealed that poverty was less strongly associated with COVID-19 risk in certain districts of southern Malawi, as illustrated in Fig. 2D. Conversely, many central and northern districts exhibited higher COVID-19 risk than expected, based solely on poverty, population density, age, and precipitation. Interestingly, high poverty levels were also observed in districts along Lake Malawi, suggesting that unmeasured factors such as inadequate sanitation or mobility patterns might be significant contributors to the elevated risk.

Wave one spanned from April to September 2020, wave two from November 2020 to February 2021, wave three from May to September 2021, and the fourth wave from November 2021 to February 2022. Relative risks ranged between 8.8 × 10*^−^*^8^ and 1.0, with an overall mean of 0.015, with this huge variability reflecting the large differences in prevalence and incidence typical of waves of an infectious disease.

For the 0-19 age group, the relative risk remained consistently low and nearly constant over time, except for an increase in Neno (southern region) between November 2020 and February 2021 (see Fig. S4 in the Supplementary Material). In contrast, other age groups showed significant variations in risk across districts and time periods.

In the age group 20-29, elevated risk was noted in Salima during the second wave as shown in Fig. S5 in the Supplementary Material. Between November 2021 and February 2022 (fourth wave), individuals in this age group experienced heightened risk in Rumphi, Mzimba, Nkhotakota, Blantyre, and Zomba. Similarly, the 30-39 age group had an increased risk in Blantyre during this period, with additional clusters identified in northern Malawi (see Fig. S6 in the Supplementary Material).

For those aged 40 and above, the highest risk occurred in Blantyre around December 2021, as illustrated in Fig. 3 and Fig. S7 in the Supplementary Material. Overall, elevated risks were most pronounced during the fourth peak (19-25 December, 2021) and, to a lesser extent, during the third peak (18-24 July, 2021). Districts with notable risks included Rumphi, Mzimba, and Nkhata Bay in the north; Nkhotakota, Lilongwe, and Salima in the central region; and Blantyre and Zomba in the south.

**Figure 3:**
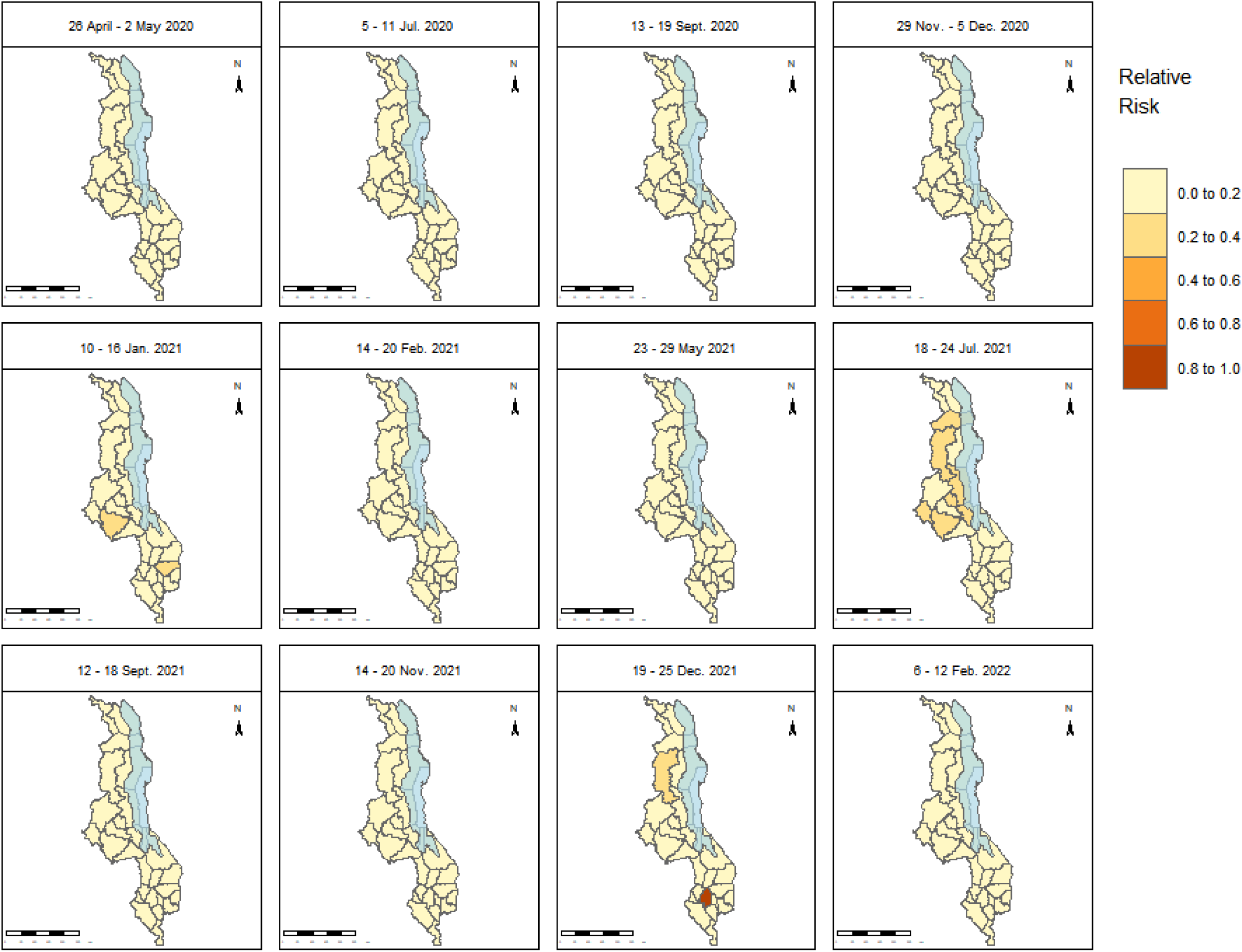
Overall relative risk maps for age group 40 − 49 in selected weeks. Highest risk was observed in Blantyre in 2021 when approaching the festive season of Christmas. The blue-shaded area in the maps is Lake Malawi. The shapefiles used to create the maps are openly available at https://data.humdata.org/dataset/cod-ab-mwi? and the link to the data licence is https://data.humdata.org/faqs/licenses.

**Figure 4:**
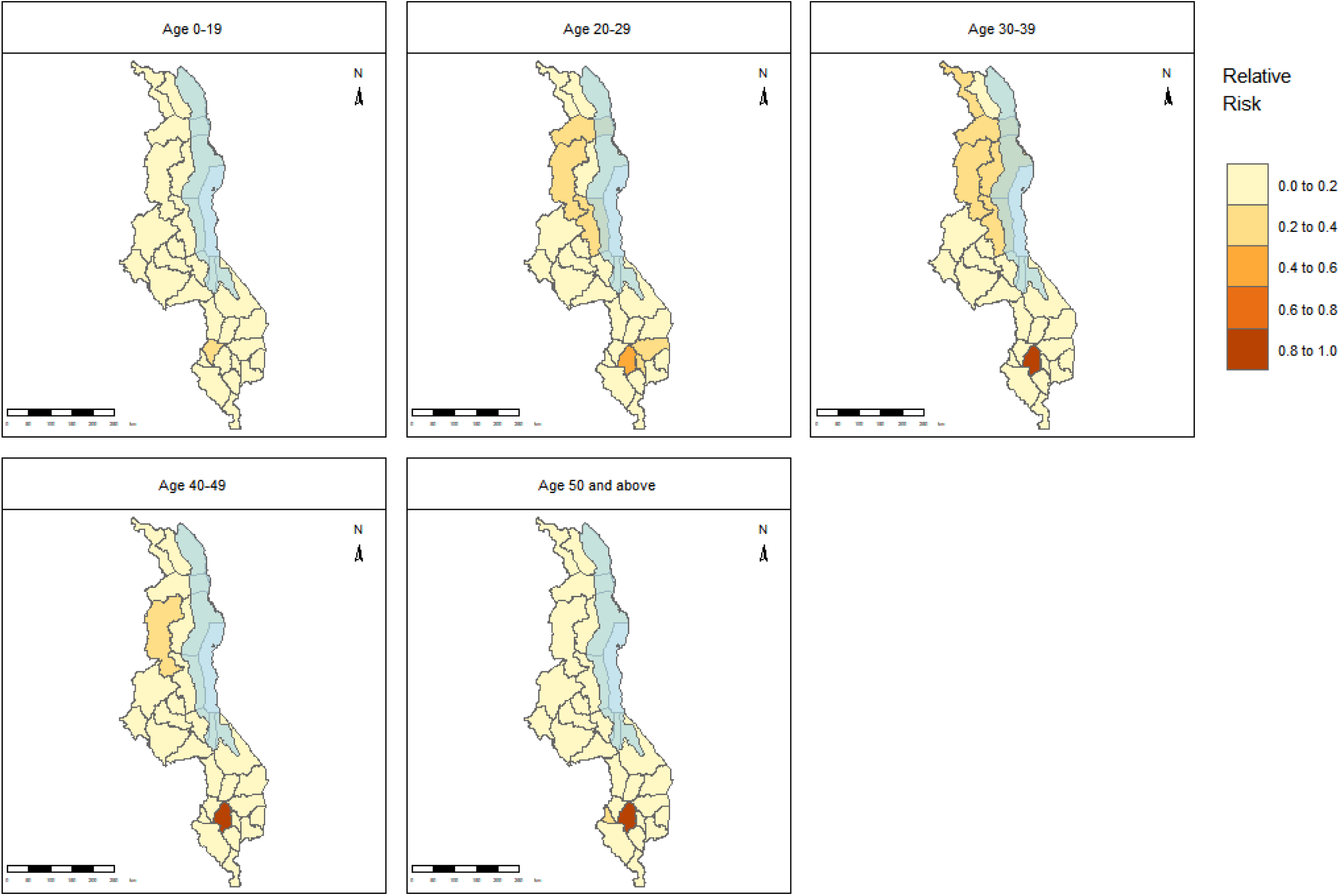
Overall relative risk maps for all age groups between 19 and 25 December 2021. Maps for all age groups during this period show an increased risk in the south, particularly in Blantyre city. The blue-shaded area in the maps is Lake Malawi. The shapefiles used to create the maps are openly available at https://data.humdata.org/dataset/cod-ab-mwi? and the link to the data licence is https://data.humdata.org/faqs/licenses.

Fig. S8 in the Supplementary Material illustrates the covariate-adjusted relative risk and the posterior spatial random effects across districts. The map emphasises significant unexplained effects, especially in southern districts such as Neno, Mwanza, Chikwawa, Nsanje, Blantyre, Thyolo, Chiradzulu, Mulanje, and Zomba, in contrast to the central and northern regions. This indicates the presence of additional unidentified factors contributing to the disease risk in these areas. Cross-border interactions with Mozambique, particularly through the port city of Beira, could contribute to disease spillover. Additionally, localized outbreaks in trade hubs like Blantyre, which has high interaction levels, might be driven by superspreading events. The presence of the highly transmissible Omicron variant, which impacted the southern region severely, could further complicate the situation. Omicron was first identified in South Africa, and the high cross-border mobility between Malawi and South Africa likely facilitated its spread into southern Malawi, where border post is located.

Temporal effects (modelled using a random walk of order 1) with 95% intervals for the 40-49 age group were further examined. The results revealed the presence of unmeasured temporal influences in the data that could not be explained by known demographic or environmental factors, such as age or precipitation. A pattern of heightened risk was observed at the end and beginning of the year, as well as mid-year. These findings suggest underlying temporal dynamics that warrant further exploration. An illustration of these results is provided in Fig. S9 in the Supplementary Material.

The variation in COVID-19 risk over time across districts for the 40-49 age group was analysed and the results delineate the four distinct waves of infection, across nearly all districts. The relative risk remained below 1 in all districts throughout the study period, except for Blantyre, indicating that the observed infection rates were lower than the baseline expectation. This consistent pattern highlights spatial and temporal similarities in how the pandemic unfolded across the districts. For a visualisation of these results, refer to Fig. S10 and Fig. S11 in the Supplementary Material.

Risk variation over time across all age groups was monitored in a few selected districts where risk was high, including the two cities, Blantyre and Lilongwe, as well as Neno. Results indicated that the 40-49 age group experienced elevated risk in Blantyre between November 2021 and February 2022. In Neno, the youngest age group was at higher risk from November 2020 to February 2021, while the oldest age group was most affected between May 2021 and February 2022. From November 2020 to February 2021 in Lilongwe, the age group 40-49 experienced the highest COVID-19 risk compared to other age groups. In contrast, from May to September 2021, the highest risk shifted to the oldest age group, 50 and above. These findings are visualised in Fig. S12 in the Supplementary Material.

Exceedance probability was used to assess how frequently the estimated risk in districts surpassed a defined threshold, enabling the identification of disease clusters and hotspots. Weekly average risk values served as the thresholds. Results for selected weeks are presented in Fig. 5. The findings revealed that risk was consistently high in the cities of Blantyre and Lilongwe. Occasionally, elevated risk was observed in Mzuzu/Mzimba and Zomba, as well as in several lakeshore districts and other areas, including Dowa, Mchinji, and Ntcheu in the central region, and Phalombe in the southern region.

**Figure 5:**
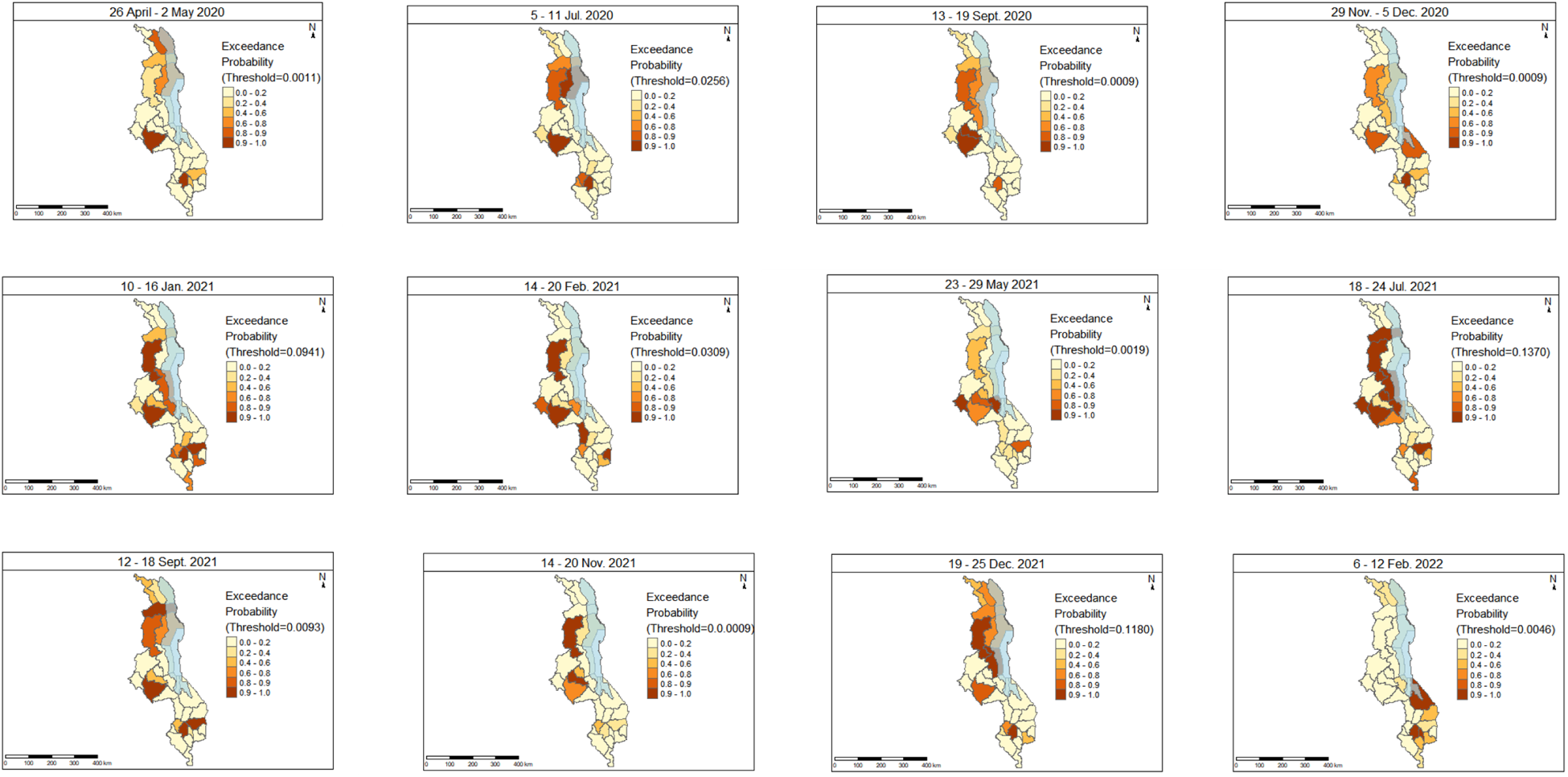
Exceedance probability for the age group 40 − 49 in selected weeks. The blue-shaded area in the maps is Lake Malawi. The shapefiles used to create the maps are openly available at https://data.humdata.org/dataset/cod-ab-mwi? and the link to the data licence is https://data.humdata.org/faqs/licenses.

## Discussion

Here we have presented a spatiotemporal model of Malawi to explore the association between age groups, poverty, population density, precipitation, and COVID-19 risk, and to identify hotspot areas. The analysis utilized data collected from 22 April 2020 to 27 March 2022. Spatiotemporal models incorporating fixed effects, spatial and temporal random effects, and interaction terms (space-time and space-time-age) were applied. Among the various models tested, the model that demonstrated superior performance, with the lowest Deviance Information Criterion (DIC) and Watanabe-Akaike Information Criterion (WAIC) scores, included covariates such as age, poverty proportion, population density, and precipitation, along with spatial effects, temporal effects, a Type 4 spatiotemporal interaction, and an independent interaction term for space, time, and age.

This study identifies significant effects of age on COVID-19 incidence, consistent with findings by Ngwira *et al.* (2021) [16] and Chinkaka *et al.* (2023) [15], although Chinkaka *et al.* (2023) [15] reported these effects as not statistically significant (*p* = 0.176). The inclusion of multiple age groups in this study reveals an interesting result: the risk is higher in the 40–49 age group than in the 50+ age group. This finding contrasts with much of the literature, where COVID-19 risk is generally associated with the oldest age groups. The elevated risk observed in the 40-49 age group may be attributed, but not limited, to several factors. Individuals in this age group may have weaker immune systems compared to younger populations (under 30), increasing their susceptibility to infection. Additionally, the 40–49 age group is often more active in the workforce, engaging in activities that may expose them to higher contact rates and transmission risks. By contrast, many individuals over 50 years may be retired and spend more time at home, potentially reducing their exposure to the virus.

This study finds a positive correlation between population density and COVID-19 risk, consistent with findings by Chinkaka *et al.* (2023) [15] and other studies conducted in Africa and abroad, including Md Iderus *et al.* (2022) [42], Nguimkeu and Tadadjeu (2021) [43], and Wong and Li (2020) [44]. However, further analysis suggests that the impact is not solely attributable to population density, age, poverty, or precipitation. Unmeasured factors, such as mobility patterns and compliance with public health measures, likely play a significant role. In contrast to Ngwira *et al.* (2021) [16], this study reveals a positive association between poverty and SARS-CoV-2 risk. This association is likely influenced by overcrowded living conditions, limited access to healthcare, and poor hygiene. Additionally, high mobility and reliance on informal work may exacerbate risk. Many individuals living in poverty depend on daily wages or informal employment, which often requires frequent travel and interactions in crowded markets or workplaces. Furthermore, poverty can limit adherence to stay-at-home orders or lockdown measures, as daily income is essential for survival. These findings highlight the multifaceted relationship between socioeconomic factors and COVID-19 risk, emphasising the need to address structural inequities and support vulnerable populations to reduce transmission

Infection rates peaked during the colder months and festive seasons. COVID-19 impacted different districts at varying times, with higher risks observed in cities and lake-shore districts. Blantyre city was the most affected. The elevated risk in cities may be attributed to the influx of tourists, as international airports are located in urban centers, facilitating the importation of cases. Similarly, the heightened risk in lake-shore areas such as Rumphi, Nkhata Bay, Nkhotakota, Salima, and Mangochi can also be explained by the presence of tourists, contributing to both imported and locally transmitted cases. The analysis of random effects further suggests that increased risk in lake-shore areas may stem from inadequate hygiene practices and limited compliance with public health interventions. In Blantyre city, the higher risk could be linked to its status as Malawi’s leading trade center, attracting large numbers of people seeking economic opportunities. This population influx likely increases crowding and mobility, amplifying transmission risks [45].

We have identified three significant limitations of this study. The first of these is that we do not have access to underlying estimates of infection prevalence and incidence independently of case finding, and hence there may be confounding with test-seeking behaviour; however, this is common to the majority of studies of infectious disease, and when studies of infection are carried out in European countries, biases due to testing were not typically large [46]. The second limitation is the absence of COVID-19 vaccination data in the model. Vaccination significantly impacts transmission and severity. Without accounting for vaccination data, the model may overestimate the risk in areas with high vaccination rates or underestimate the risk in districts with low vaccination uptake. Furthermore, the spatial and temporal variability of vaccine rollouts, influenced by socio-economic and political factors, adds complexity that the model does not account for. Including vaccination data in future models would provide a more accurate and comprehensive understanding of COVID-19 dynamics. The third notable limitation of this study is the use of the average relative risk for the given week as the threshold for calculating exceedance probability. This approach is suboptimal, as the threshold should ideally be derived from expert judgment or established standards to ensure greater accuracy and relevance.

This work opens several avenues for future exploration and enhancement. The current analysis employs a Conditional Autoregressive (CAR) structure, which is based on the contiguity of districts. While effective, it may be beneficial to explore alternative spatial structures, such as Gaussian Random Fields (GRFs), which are more flexible and allow for smoother modelling of spatial relationships based on distance rather than strict adjacency [47, 48]. GRFs can better capture subtle spatial dependencies and provide a richer representation of spatial variation. Furthermore, we can also incorporate a more advanced mathematical modelling approach that integrates both deterministic and stochastic elements, providing a comprehensive framework to better capture the complex dynamics of COVID-19 transmission. This approach will enhance our understanding of how different factors influence the spread of COVID-19 over time and across different regions [49].

## Conclusion

This study identifies significant effects of space, time, age, population density, and poverty on COVID-19 transmission in Malawi, while the effect of precipitation was not significant. Notably, the risk was highest during the colder months (June and July), the festive season (December), and January. Urban areas and districts along Lake Malawi were more affected than other areas, with the 40–49 age group being at particularly high risk. Given these findings, if COVID-19 resurges or if similar infectious diseases with comparable characteristics emerge in the future, priority should be given to vaccinating the working population in urban areas and tourist centres during cold months and festive seasons. Additionally, the government must ensure that individuals in these high-risk areas comply with public health interventions and have access to adequate healthcare services.

## Data Availability

Links to all data used in this study are provided in the manuscript.

## Supporting information

*S1.* Supplementary Material

### Use of AI for Language Enhancement

Artificial intelligence (AI)-assisted tools were used to enhance the readability and grammatical accuracy of this manuscript. However, all intellectual content, interpretations, and conclusions remain the responsibility of the authors.

## Declaration of Competing Interest

None

## 4. Ethics approval

This study was approved by the National Commission for Science and Technology, Malawi (Approval No: P.02/23/733).

## Funding

MKA was supported by the Schlumberger Foundation–Faculty for the Future. OJ was supported by the Wellcome Trust (228264/Z/23/Z). IH and TH were supported by the Wellcome Trust (227438/Z/23/Z) and the UKRI Impact Acceleration Account (IAA 386).

## Notes

### Competing Interest Statement

The authors have declared no competing interest.

### Funding Statement

This study was funded by the Schlumberger Foundation-Faculty for the Future and the Wellcome Trust.

### Author Declarations

Ethics committee of the National Commission for Science and Technology, Malawi (Approval No: P.02/23/733) gave ethical approval for this work.

