## Supplementary Material for "Spatiotemporal Analysis of COVID-19 Risk in Malawi: The Impact of Age, Poverty, Population Density, and Environmental Factors"

#### S1. Methods

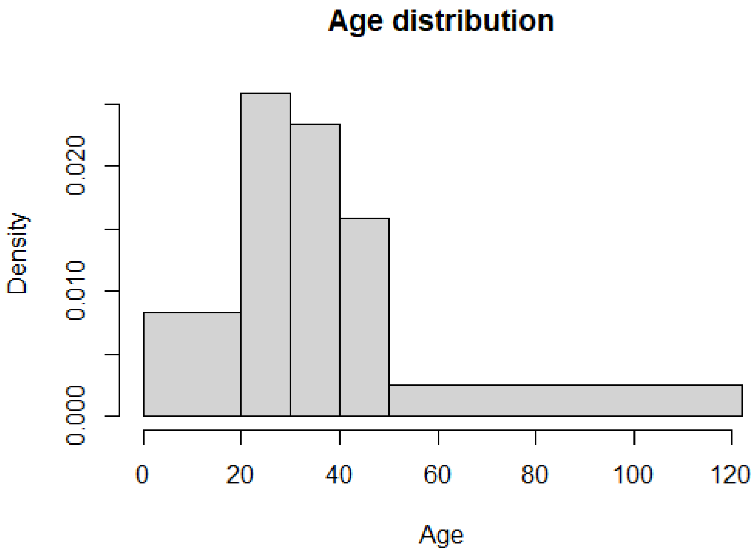


Figure S1: **Age histogram**

*S1.1. Prior specification*

The structured spatial random effects follows a normal distribution with mean; random effects in the neighbourhood and variance inversely proportional to the number of neighbours.

$$u_i|\mathbf{u_{-i}} \sim N\left(\overline{u}_{\sigma_i}, \frac{1}{\tau_u n_{\sigma_i}}\right), \tag{1}$$

where $\sigma_i$ = set of neighbours, $n_{\sigma_i}$ = number of neighbours, $\tau_u = \frac{1}{\sigma_u^2}$ and

$$\overline{u}_{\sigma_i} = \frac{1}{n_{\sigma_i}} \sum_{r \in \sigma_i} u_r.$$

Unstructured spatial effects, $v_i$ follow a normal distribution;

$$v_i \sim N\left(0, \frac{1}{\tau_v}\right). \quad (2)$$

Three structures for temporal effects, $u_j$ are considered including:

(i) linear time trend $u_j \sim N(\beta t_j, \tau)$, where $t_j$ is time associated with $j^{th}$ week.

(ii) random walk 1 (rw1): $u_j \sim N(u_{j-1}, \tau)$ where rw1 prior distribution is

$$\pi(u|\sigma_u^2) \propto exp\left(-\frac{1}{2\sigma_u^2}\sum_{j=2}^{J}(u_j - u_{j-1})^2\right)$$

(iii) random walk 2 (rw2): $u_j \sim N(2u_{j-1} + u_{j-2}, \tau)$ where rw2 is

$$\pi(u|\sigma_u^2) \propto exp\left(-\frac{1}{2\sigma_u^2}\sum_{j=2}^{J}(u_j - 2u_{j-1} + u_{j-2})^2\right)$$

The unstructured temporal effect is given as $v_j \sim N(0, \sigma_v^2)$

The space-time interaction term, $\phi_{ij}$, is classified into four types (Fattah and Rue, 2022), derived from all possible combinations of structured and unstructured spatial and temporal effects (Knorr-Held, 2000) as follows:

1. Type I interaction follows a normal distribution with mean 0 and variance $\sigma_\phi^2$; $\phi_{i,j} \sim N(0, \sigma_\phi^2)$.

$$\pi(\phi|\tau_\phi) \propto \exp\left(-\frac{\tau_\phi}{2}\sum_{i=1}^{28}\sum_{j=1}^{105}\phi_{ij}^2\right),$$

where $\tau$ is a precision parameter.

2. Type II interaction involves structured temporal effects and unstructured spatial effects. In this case, $\phi_i = (\phi_{i1}, \phi_{i2}, \cdots, \phi_{i105})^T$, where each area follows a random walk, and neighbouring areas have independent temporal correlations.

$$\pi(\phi|\tau_\phi) \propto \exp\left(-\frac{\tau_\phi}{2}\sum_{i=1}^{28}\sum_{j=1}^{105}(\phi_{ij} - \phi_{ij-1})^2\right).$$

3. Type III interaction, where each $\phi_j = (\phi_{1j}, \phi_{2j}, \cdots, \phi_{28j})^T$ follows an independent intrinsic auto-regression.

$$\pi(\phi|\tau_\phi) \propto \exp\left(-\frac{\tau_\phi}{2}\sum_{j=1}^{105}\sum_{i\sim l}(\phi_{ij} - \phi_{lj})^2\right),$$

where $l$ is the number of covariates.

4. Type IV interaction, where;

$$\pi(\phi|\tau_\phi) \propto \exp\left(-\frac{\tau_\phi}{2}\sum_{t=2}^{T}\sum_{j\sim i}(\phi_{jt} - \phi_{it} - \phi_{jt-1} - \phi_{it-1})^2\right).$$

The four interaction terms are extended by the space-time-age interaction, $\psi_{ijk}$, which follows a normal distribution with a mean of 0 and a variance of $\sigma^2_\psi$. The random effects of poverty and population density are modelled using random walk 1.

*S1.2. Inference*

*S1.2.1. Integrated Nested Laplace Approximations (INLA)*

Analysis was done in `R` using `INLA` package Rue et al. (2009). INLA can be installed from `www.r-inla.org`. INLA provides an approximate Bayesian inference framework for latent Gaussian models. This approach employs analytical approximations and numerical integration to obtain the posterior distribution of parameters. Compared to Markov Chain Monte Carlo (MCMC), INLA is faster and efficiently handles large datasets. Observations in latent Gaussian models follow a distribution from the exponential family;

$$y_i|\mathbf{z}, \theta \sim \pi(y_i|z_i, \theta)$$

$i = 1, 2, ..., n$. The parameters, $\mathbf{z}$ follow a Gaussian distribution;

$$\mathbf{z}|\theta \sim N(\mu(\theta), \mathbf{Q}(\theta)^{-1})$$

The hyper-parameters, $\theta$ follow a default log-gamma distribution for INLA, $\log -\Gamma(1, 0.0005)$. So, the linear predictor is the sum of an intercept, covariates and random effects.

$$\eta_i = \alpha + \sum_{k=1}^{n_\beta}\beta_k Z_{ki} + \sum_{j=1}^{n_f} f^{(j)}(u_{ji})$$

INLA computes posterior marginals of latent gaussian field and hyper-parameters;

$$\pi(z_i|\mathbf{y}) = \int \pi(z_i|\theta, \mathbf{y})\pi(\theta|\mathbf{y})d\theta$$

And

$$\pi(\theta_{\mathbf{j}}|\mathbf{y}) = \int \pi(\theta|\mathbf{y})d\theta_{-j}$$

Integrands are then approximated using Laplace approximations;

$$\tilde{\pi}(z_i|\mathbf{y}) = \int \tilde{\pi}(z_i|\theta, \mathbf{y})\tilde{\pi}(\theta|\mathbf{y})d\theta \tag{3}$$

And

$$\tilde{\pi}(\theta_{\mathbf{j}}|\mathbf{y}) = \int \tilde{\pi}(\theta|\mathbf{y})d\theta_{-j} \tag{4}$$

Numerically integrating 5 and 6 leads to;

$$\tilde{\pi}(z_i|\mathbf{y}) = \sum_k \tilde{\pi}(z_i|\theta_{\mathbf{k}}, \mathbf{y})\tilde{\pi}(\theta_{\mathbf{k}}|\mathbf{y}) \times \triangle_k$$

And

$$\tilde{\pi}(\theta_{\mathbf{j}}|\mathbf{y}) = \sum_l \tilde{\pi}(\theta^*_{\mathbf{l}}|\mathbf{y}) \times \triangle^*_l$$

where $\triangle_k$ and $\triangle^*_l$ are area weight corresponding to $\theta_k$ and $\theta^*_l$ respectively.

*S1.2.2. Deviance Information Criterion (DIC) and Watanabe-Akaike Information Criterion (WAIC)*

Selection of the best model is based on the scores of deviance information criterion (DIC) Spiegelhalter et al. (2002) and the Watanabe-Akaike information criterion (WAIC) (Watanabe and Opper, 2010; Watanabe, 2021). DIC assesses model complexity and goodness of fit. The equation for DIC, which is the sum of the measure of model fit and the effective number of parameters, is provided below.

$$\text{DIC} = -2\log L\big(Y\big|\hat{\theta}_{\text{Bayes}}\big) + 2P_{\text{DIC}}$$

where $P_{\text{DIC}}$ is the difference between the posterior mean of the deviance and the deviance at the posterior means of the parameters of interest (Spiegelhalter et al., 2002). As the models are Bayesian, the WAIC is also utilized, as it is specifically designed to evaluate and measure predictive accuracy in Bayesian frameworks. The equation for WAIC is provided below:

$$\text{WAIC} = -2(\text{lppd} - P_{\text{WAIC}})$$

where lppd is the log pointwise predictive density while $P_{\text{WAIC}}$ is the effective number of parameters (Watanabe and Opper, 2010). The model with the least DIC, WAIC and effective number of parameters for WAIC is considered the best.

### Results

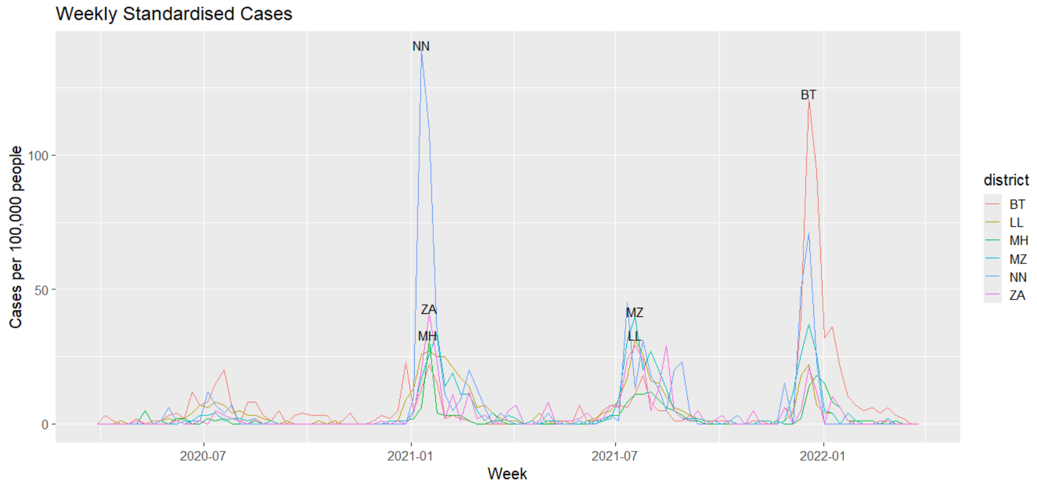


Figure S2: **District standardised case counts.** Neno and Blantyre experienced high risk during the second and fourth waves, respectively.

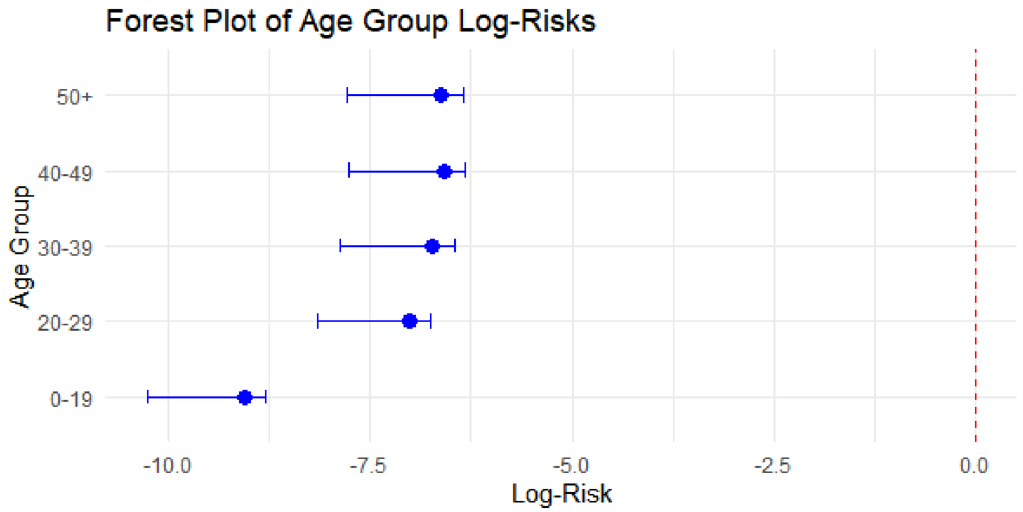


Figure S3: **Forest plot for age groups:** The Log-Risks of all age groups is below zero, which implies that the risk in the groups is lower than the baseline assumed by the model, usually 1.

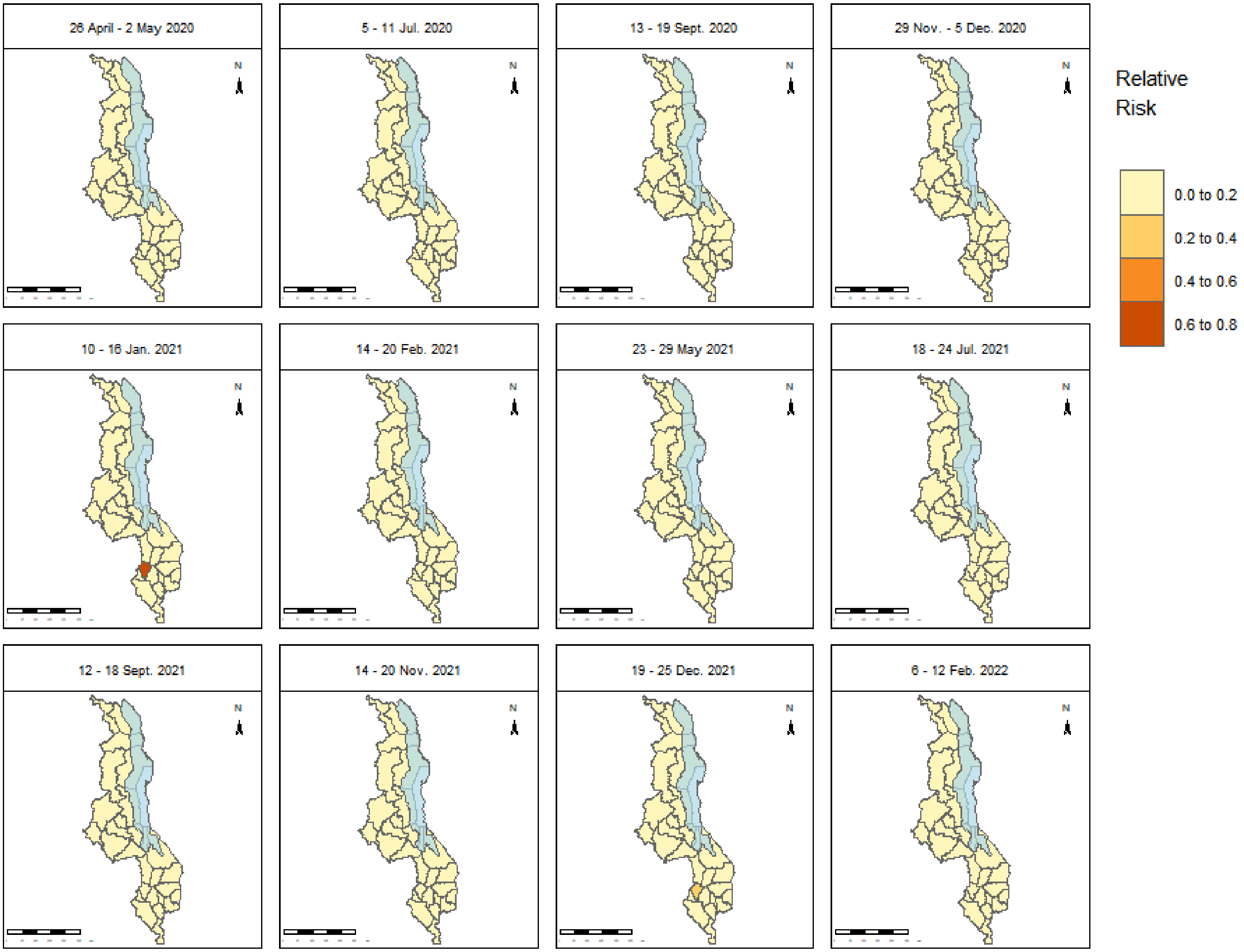


Figure S4: **Overall relative risk maps for** $0-19$ **in selected weeks.** Risk during the study period was generally low in all districts, except for Neno district in the southern region, during January 10–16, 2021, and December 19–25, 2022. The blue-shaded area in the maps is Lake Malawi.The shapefiles used to create the maps are openly available at https://data.humdata.org/dataset/cod-ab-mwi? and the link to the data licence is https://data.humdata.org/faqs/licenses.

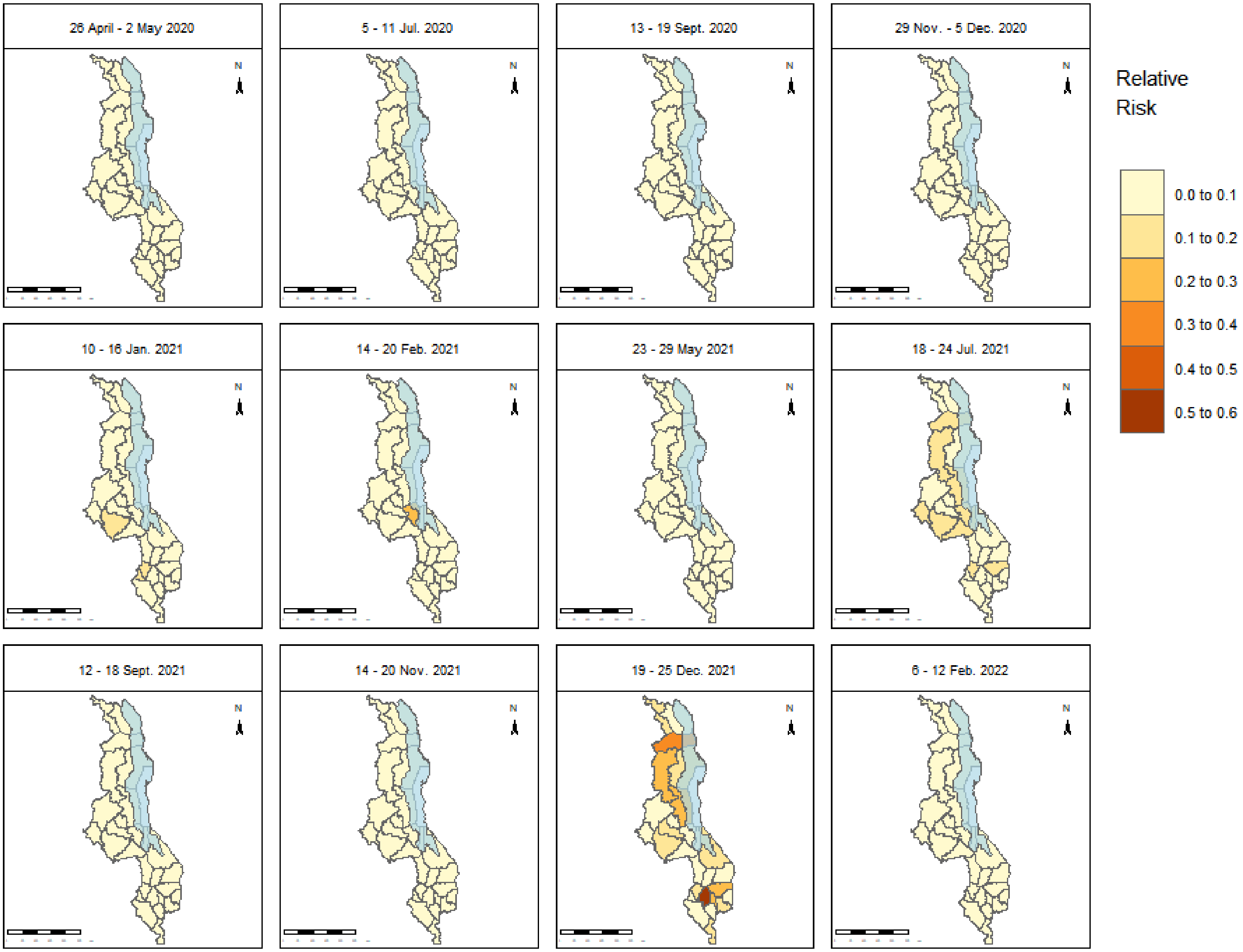


Figure S5: **Overall relative risk maps for age group** $20 - 29$ **in selected weeks.** Higher risk was observed across the country especially between 19 and 25 December 2021. Blantyre in the south had highest risk. The blue-shaded area in the maps is Lake Malawi. The shapefiles used to create the maps are openly available at https://data.humdata.org/dataset/cod-ab-mwi? and the link to the data licence is https://data.humdata.org/faqs/licenses.

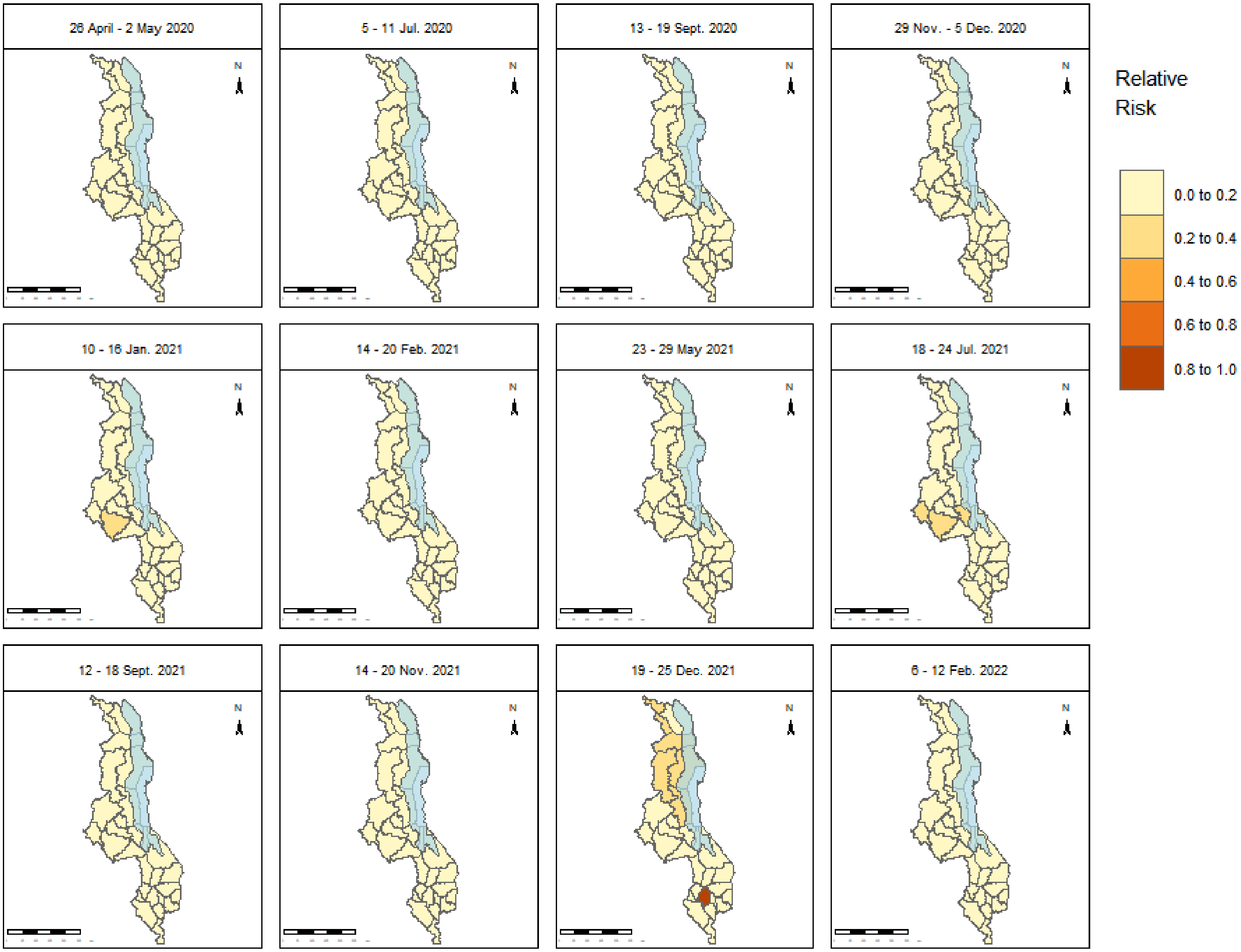


Figure S6: **Overall relative risk maps for age group** $30 - 39$ **in selected weeks.** From 18 to 24 July 2021, risk was higher in the central part, the capital city Lilongwe, Mchinji and Salima, than the rest of the areas. Between 19 and 25 December 2021, risk was high in most of the northern areas but extremely high in Blantyre in the south. The blue-shaded area in the maps is Lake Malawi. The shapefiles used to create the maps are openly available at https://data.humdata.org/dataset/cod-ab-mwi? and the link to the data licence is https://data.humdata.org/faqs/licenses.

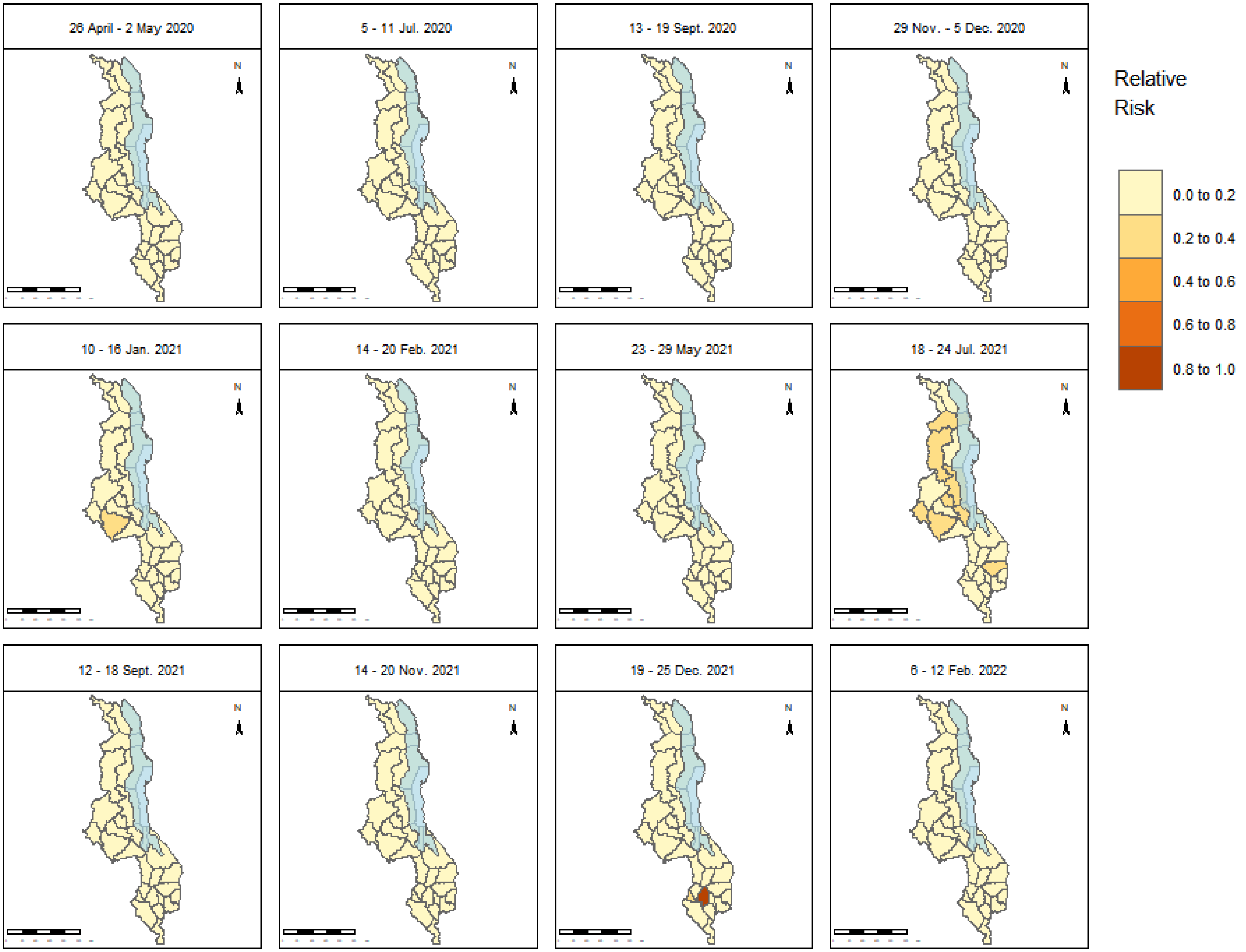


Figure S7: **Overall relative risk maps for age group** 50+ **in selected weeks.** High risk was observed from 18 to 24 July 2021 across the country in areas such as Rumphi, Mzimba, Nkhotakota, Salima, Lilongwe, Mchinji and Zomba. The blue-shaded area in the maps is Lake Malawi. The shapefiles used to create the maps are openly available at https://data.humdata.org/dataset/cod-ab-mwi? and the link to the data licence is https://data.humdata.org/faqs/licenses.

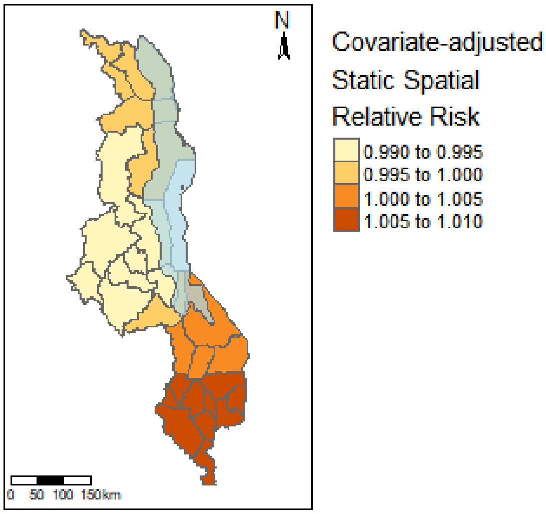


Figure S8: **Map showing the covariate-adjusted relative risk.** The blue-shaded area is Lake Malawi. The shapefiles used to create the map are openly available at `https://data.humdata.org/dataset/cod-ab-mwi?` and the link to the data licence is `https://data.humdata.org/faqs/licenses`.

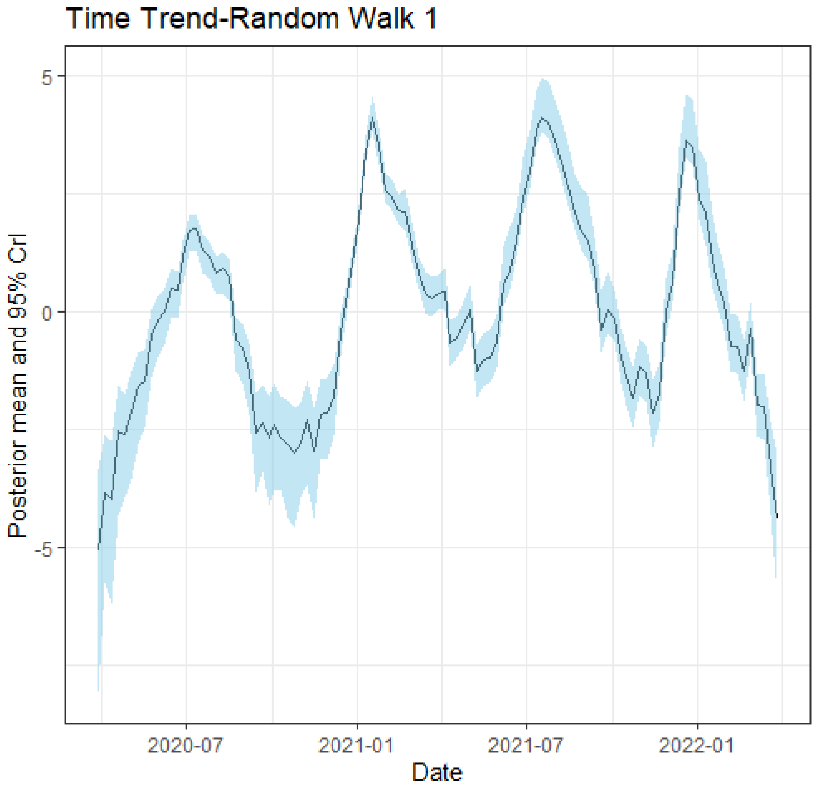


Figure S9: Temporal random effects for age group (40 – 49). Random walk 1.

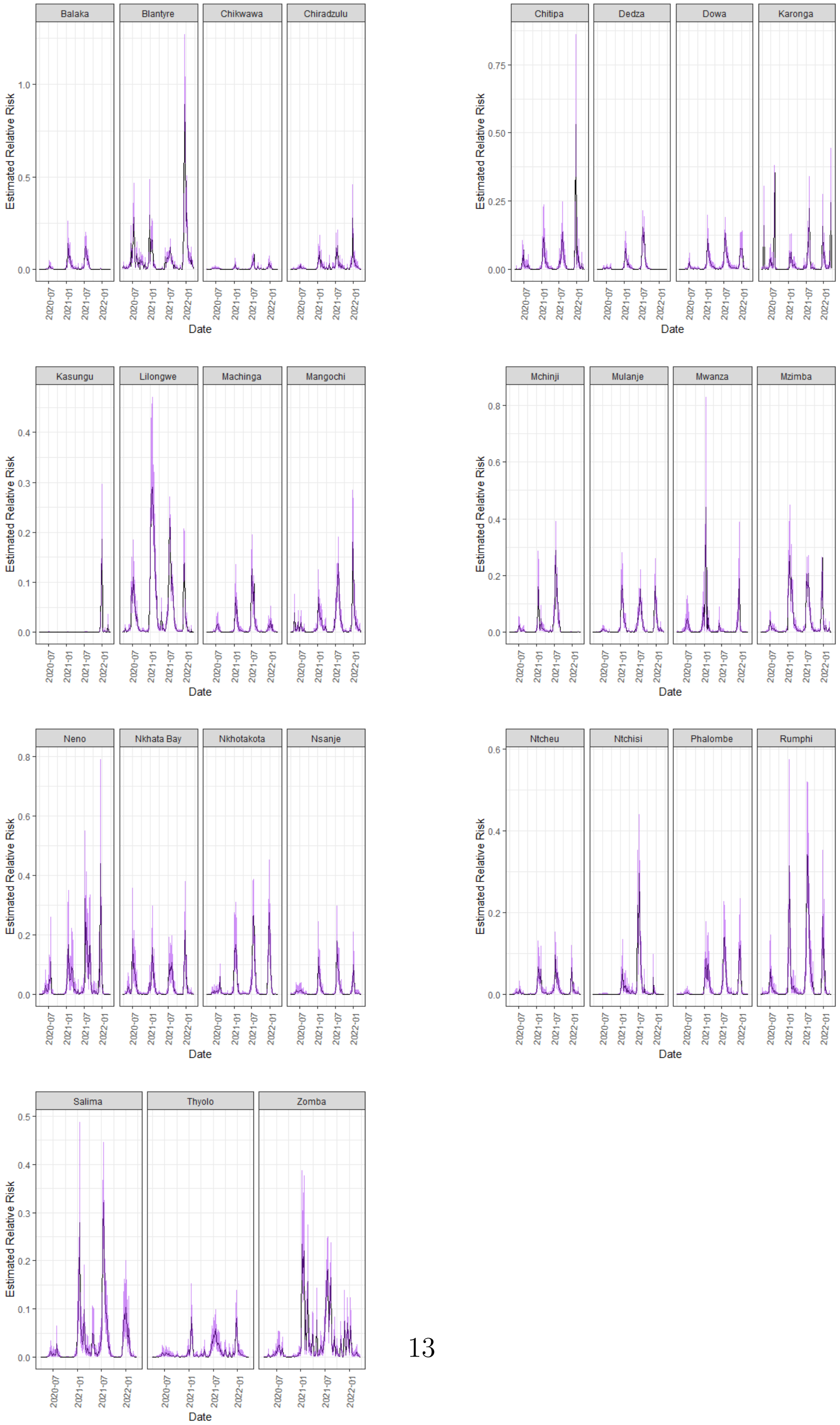


Figure S10: Variation of relative risk and 95% CI over time in districts for age group $40-49$.

Fig.S11 illustrates the variation in COVID-19 risk over time for the 40–49 age group across districts in a single plot. Notably, there are elevated risk levels in Blantyre, although they remain below 1. Interestingly, these findings align closely with the high-risk areas identified in Fig.S2, reinforcing the consistency of the results.

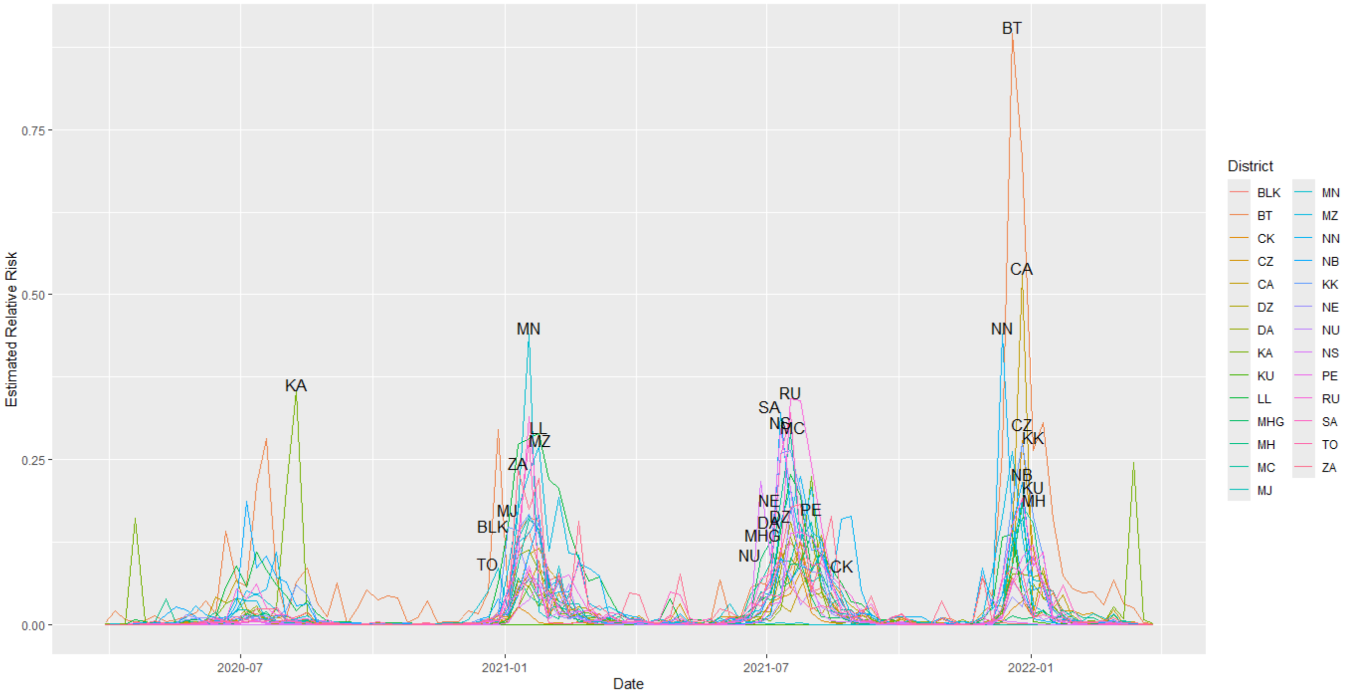


Figure S11: **Variation of relative risk for age group** $40-49$ **in all districts.**

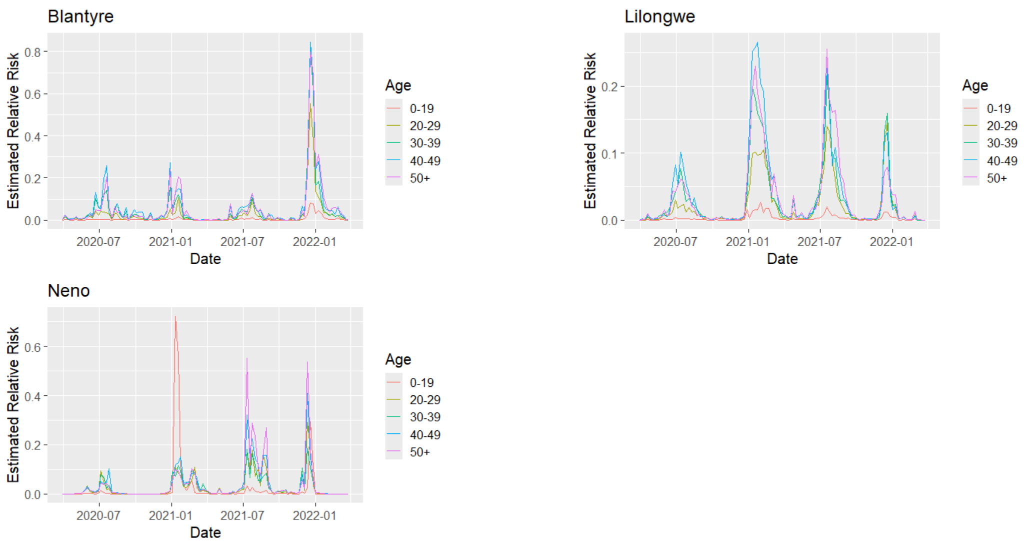


Figure S12: Variation of relative risk over time in age groups for selected districts
